# High incidence of asymptomatic SARS-CoV-2 infection, Chongqing, China

**DOI:** 10.1101/2020.03.16.20037259

**Authors:** Yang Tao, Panke Cheng, Wen Chen, Peng Wan, Yaokai Chen, Guodan Yuan, Junjie Chen, Da Huo, Ge Guan, Dayu Sun, Ju Tan, Guanyuan Yang, Wen Zeng, Chuhong Zhu

## Abstract

**Background:** SARS-CoV-2 has been a global pandemic, but the emergence of asymptomatic patients has caused difficulties in the prevention of the epidemic. Therefore, it is significant to understand the epidemiological characteristics of asymptomatic patients with SARS-CoV-2 infection.

**Methods:** In this single-center, retrospective and observational study, we collected data from 167 patients with SARS-CoV-2 infection treated in Chongqing Public Health Medical Center (Chongqing, China) from January to March 2020. The epidemiological characteristics and variable of these patients were collected and analyzed.

**Findings:** 82.04% of the SARS-CoV-2 infected patients had a travel history in Wuhan or a history of contact with returnees from Wuhan, showing typical characteristics of imported cases, and the proportion of severe Covid-19 patients was 13.2%, of which 59% were imported from Wuhan. For the patients who was returnees from Wuhan, 18.1% was asymptomatic patients. In different infection periods, compared with the proportion after 1/31/2020, the proportion of asymptomatic patient among SARS-CoV-2 infected patient was higher(19% *VS* 1.5%). In different age groups, the proportion of asymptomatic patient was the highest(28.6%) in children group under 14, next in elder group over 70 (27.3%). Compared with mild and common Covid-19 patients, the mean latency of asymptomatic was longer (11.25 days *VS* 8.86 days), but the hospital length of stay was shorter (14.3 days *VS* 16.96 days).

**Conclusion:** The SARS-CoV-2 prevention needs to focus on the screening of asymptomatic patients in the community with a history of contact with the imported population, especially for children and the elderly population.

## Introduction

WHO has announced on March 11 in Switzerland that SARS-CoV-2 has the characteristics of a global pandemic ^1^. As of March 15, 2020, China has accumulated 81,048 cases of SARS-CoV-2 infection and a mortality rate of 3.95 %, also, there is a total of 64322 cases expect China, with a mortality rate of 3.51%^1,2^. The proportion of asymptomatic patients has gradually increased ^3^. Asymptomatic patients are still highly contagious, paying special attention to the epidemiological characteristics of asymptomatic patients is of great significance for the control of the epidemic.

Wuhan, Hubei was the first outbreak location in China^4^. Chongqing is a province adjacent to Hubei, as of March 15, had 576 cases of SARS-CoV-2 infection, and a mortality rate of 1.04%^5^. However, the epidemiological characteristics of SARS-CoV-2 infected patients in the input area are not clear at present, especially for the asymptomatic patients. This article takes Chongqing, China, as a representative area of importation case, analyzes the case data of SARS-CoV-2 infected patients, especially for the asymptomatic patients, to provide clinical reference for the prevention of SARS-CoV-2 infection.

## Materials and Methods

### Patients

The clinical data of 167 SARS-CoV-2 infected patients treated in Chongqing Public Health Medical Center from January 2020 to March 2020 were collected. COVID-19 is diagnosed according to the WHO Interim Guidelines^6^. The severity of COVID-19 was defined according to the American Thoracic Society’s Community Acquired Pneumonia Guidelines^7^. A confirmed case of SARS-CoV-2 infection was defined as a positive result on RT-PCR assay of nasal and pharyngeal swab specimens. This study was approved by the Ethics Committee of Chongqing Public Health Medical Treatment Center (NO.2020-018-01-KY), informed consent was allowed to be waived.

### Data collection

We reviewed and analyzed the clinical files, radiologic and laboratory results of all 167 patients. All data are completed through independent survey forms to ensure that they do not interfere with each other. At the same time, three investigators complete the data collection and management to ensure the independence, completeness and authenticity of the data.

### Radiologic and laboratory detection

The detection of SARS-CoV-2 was determined based on new coronavirus pneumonia diagnosis and treatment plan developed by the National Health Committee of the People’s Republic of China. All the patients performed the RT-PCR assay of nasal and pharyngeal swab specimens for SARS-CoV-2 detection^8^. Radiologic and laboratory detection of all patients were completed according to normal inspection procedures.

### Statistical Analysis

Data in the full text are expressed as median, mean ± SD, or percentage. The missing data are not counted. Statistical analysis in this study were conducted with SPSS 25.0.

## Results

### 1. Epidemiological characteristics of SARS-CoV-2 infected patients in Chongqing

There were 22 patients with severe COVID-19, and the proportion of severe COVID-19 in Recently visited Wuhan was highest (24.1%), accounting for 59% of the total number of severe Covid-19. 20 asymptomatic patients with SARS-CoV-2 infection, 2 cases (3.7%) of the returnees from Wuhan; and the highest proportion (15 cases, 18.1%) of those who had contacted with returnees from Wuhan, accounting for 75% of the total number of asymptomatic patients. The median age of 167 SARS-CoV-2 infected patients was 46 years old, COVID-19 patients was 55 years old, and asymptomatic patients was 47 years old. Overall, the proportion of severe COVID-19 was 13.2%; 36.4% of the SARS-CoV-2 infected patients over 70 years old was severe COVID-19 patients. It is worth noting that 28.6% of them who were infected with SARS-CoV-2 in children were asymptomatic patients, and 27.3% of them who were over 70 years old, suggesting that the population of children and elder may be the main population with asymptomatic infection. Before 1/31/2020, the proportion of severe COVID-19 was significantly higher than the later (17.9% *vs*. 10%). However, the proportion of asymptomatic patients after 1/31/2020 was significantly higher than before (19% *vs*. 1.5%). Due to the closure of Wuhan, Hubei Province on January 23,2020, indicating that a large number of asymptomatic patients may not be directly imported cases. As shown in Table 1.

**Table 1.**
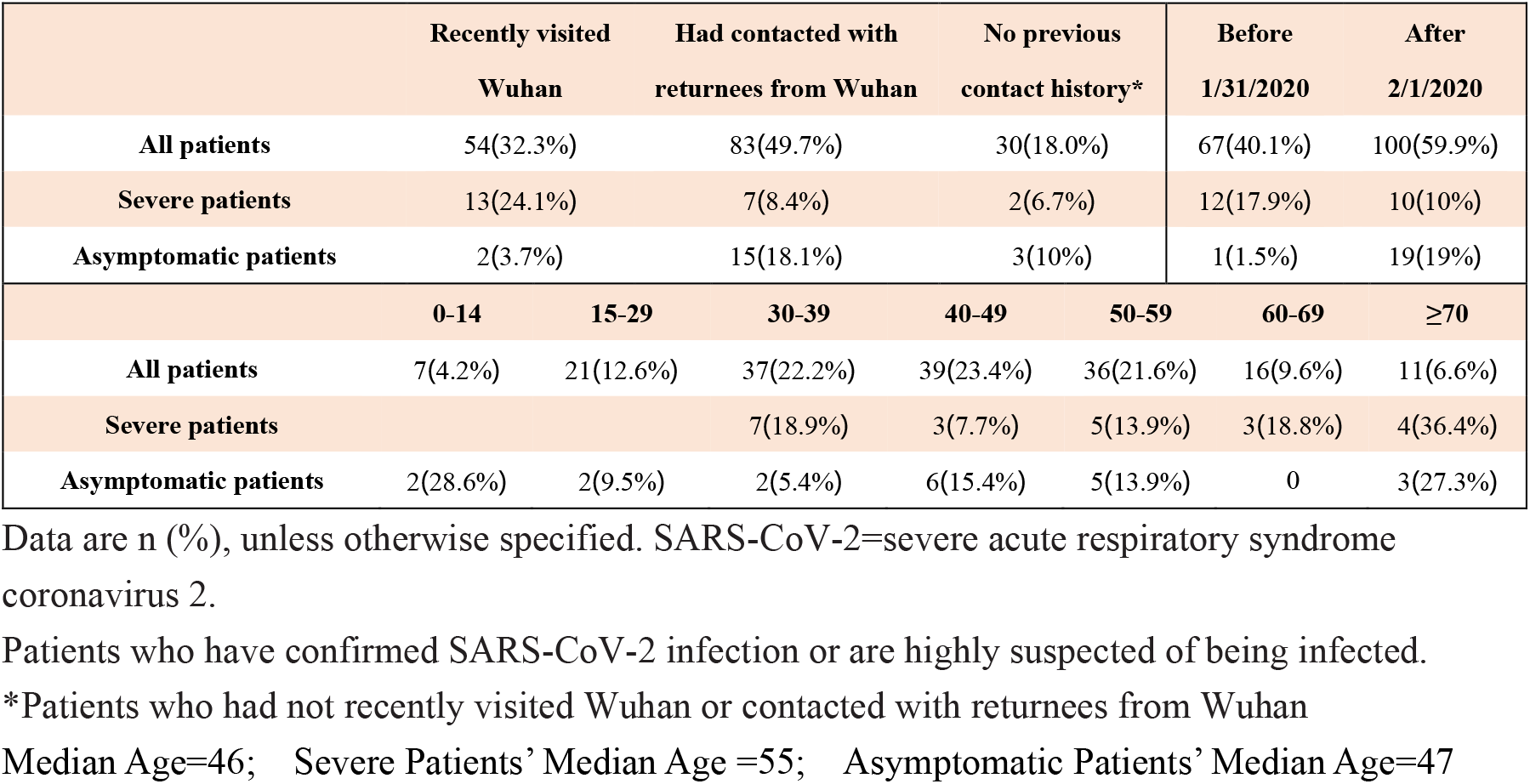
Epidemiological characteristics of SARS-CoV-2 infected patients

### 2. Clinical characteristics of severe covid-19 in Chongqing

59.09% of severe COVID-19 patients had a history of travel to Wuhan, which was a typical imported case. The median age of 22 severe COVID-19 patients was 55 years old. For the severe COVID-19 patients, the changes of clinical characteristics, radiologic and laboratory indexes were consistent with the previous reports; only 3 (13.64%) severe patients have comorbidities of liver dysfunction, lower than the previous reports(25.43%)^9^. See Table 2 for the detailed data.

**Table 2.** Variable of severe patients at different ages

|  | 30-39 (n=7) | 40-49(n=3) | 50-59(n=5) | 60-69(n=3) | ≥70(n=4) |
| --- | --- | --- | --- | --- | --- |
| All patients before 1/31/2020 | 5(71.4%) | 1(33.3%) | 3(60%) | 1(33.3%) | 2(50%) |
| **Exposure history** |  |  |  |  |  |
| Recently visited Wuhan | 5(71.4%) | 2(66.7%) | 4(80%) | 0 | 2(50%) |
| Contact with Wuhan residents | 2(28.6%) | 1(33.3%) | 1(20%) | 1(33.3%) | 2(50%) |
| Smoking history | 2(28.6%) | 0 | 1(20%) | 2(66.7%) | 1(25%) |
| Mean latency (days) | 9.1 | 8.3 | 12.3 | -- | 10.3 |
| **Clinical symptoms** |  |  |  |  |  |
| Fever | 7(100%) | 2(66.7%) | 4(80%) | 1(33.3%) | 2(50%) |
| Cough | 5(71.4%) | 2(66.7%) | 3(60%) | 2(66.7%) | 3(75%) |
| **Shortness of breath** | 1(14.3%) | 1(33.3%) | 3(60%) | 2(66.7%) | 4(100%) |
| **Fatigue** | 4(57.1%) | 0 | 1(20%) | 3(100%) | 2(50%) |
| **Comorbidities** |  |  |  |  |  |
| ARDS | 0 | 0 | 0 | 0 | 0 |
| Acute kidney injury | 0 | 0 | 0 | 0 | 0 |
| Myocardial injury | 0 | 0 | 0 | 0 | 0 |
| Liver dysfunction | 1(14.3%) | 0 | 1(20%) | 0 | 1(25%) |
| Urinary infection | 0 | 0 | 0 | 0 | 0 |
| Digestive system damage | 0 | 0 | 0 | 0 | 0 |
| **Abnormalities on chest CT** |  |  |  |  |  |
| Ground-glass opacity | 6(85.7%) | 1(33.3%) | 2(40%) | 0 | 2(50%) |
| Bilateral patchy shadowing | 3(42.9%) | 2(66.7%) | 1(33.3%) | 0 | 1(25%) |
| No obvious abnormalities | 0 | 0 | 0 | 0 | 0 |
| **Laboratory findings** |  |  |  |  |  |
| White-cell count Median (IQR) | 5.29(4.1-8.2) | 3.3(3.26-3.34) | 5.17(3.2-6.07) | 6.34(5.43-6.68) | 3.05(2.25-3.85) |
| Neutrophil Percentage median(IQR) | 60.3%<br>(57.7-75.6%) | 60.7%<br>(58-77.5%) | 72.3%<br>(63.9-75.1%) | 82.7%<br>(70.9-87.1%) | 69.8%<br>(64.2-77.6%) |
| Lymphocyte count Median (IQR) | 2.19(0.67-2.97) | 0.855(0.75-0.96) | 0.78(0.75-1.07) | 0.69(0.63-1.43) | 0.5(0.4-0.97) |
| T cell detection |  |  |  |  |  |
| CD4+ T cell count | 399(85-881) | 393(301-1083) | 167(104-226) | 117(101-356) | 176(159-230) |
| CD8+ T cell count | 342(62-692) | 304(151-812) | 235(154-316) | 121(74-404) | 111(96-141) |
| CD4+/CD8+ | 1.18(0.66-1.64) | 1.33(1.29-1.99) | 0.69(0.67-0.71) | 0.88(0.83-1.59) | 1.44(1.25-2.41) |
| Platelet count, Median (IQR) | 205.5(133-503) | 186(174-233) | 77(68-86) | 158(131-356) | 130.5(121-140) |
| Elevated C-reactive protein<br>(>10 mg/L) | 3(42.9%) | 1(33.3%) | 1(20%) | 2(66.7%) | 4(100%) |
| Elevated Procalcitonin (>0.5ng/ml) | 3(42.9%) | 1(33.3%) | 2(20%) | 1(33.3%) | 3(75%) |
| **Clinical outcome** |  |  |  |  |  |
| Recovery | 7(100%) | 3(100%) | 5(100%) | 3(100%) | 4(100%) |
| Hospital length of stay (days) | 18.3 | 17 | 26.5 | 15.7 | 21.25 |
| Death | 0 | 0 | 0 | 0 | 0 |

Data are median (IQR), n (%) or mean (SD). Some patients lack certain tests, or some patients' mean latency is unknown. White blood cell count ( $\times 10^9$ cells per L). Lymphocyte count ( $\times 10^9$ cells per L). Platelet count, $\times 10^9$ per L. ARDS=acute respiratory distress syndrome. Patients means people who have confirmed SARS-CoV-2 infection or are highly suspected of being infected.

### 3. Clinical characteristics of asymptomatic patients in Chongqing

For the asymptomatic patients, compared with mild and common Covid-19 patients, the mean latency of 50-59 age group (7 days *vs*. 9.6 days) and children group (9 days *vs*. 11.7 days) were shorter, and other age group was significantly longer, especially in the 30-39 age group (20.5 days *vs*. 7.2 days), existence C-reactive protein (3 cases, 15% *vs*. 33 cases, 26.4%) and procalcitonin (2 cases, 10% *vs*. 11 cases, 8.8%) increase. Most of the asymptomatic patients did not show typical and abnormal clinical characteristics, radiologic and laboratory indexes, only 1 (5%) had myocardial injury. In particular, some of the asymptomatic patients also had atypical changes in the radiologic indexes, which were not the diagnostic criterion of Covid-19, such as bronchiectasis, general inflammatory changes in the lung, etc. For the asymptomatic patients over the age of 70, there were 2 (66.67%) patients with ground-glass opacity in radiologic examination. All the SARS-CoV-2 infected patients were treated according to the new coronavirus pneumonia diagnosis and treatment plan developed by the National Health Committee of the People’s Republic of China. At present, all the patients have recovered. For the asymptomatic patients, compared with the mild and common Covid-19 patient, the hospital length of stay in children group was prolonged (16.5 days vs 9.8 days). See Table 3 and Table S1 for the detailed data.

**Table 3.** Variable of asymptomatic patients at different ages

|  | 0-14(n=2) | 15-29(n=2) | 30-39(n=2) | 40-49(n=6) | 50-59(n=5) | ≥70(n=3) |
| --- | --- | --- | --- | --- | --- | --- |
| **All patients before 2020.1.31** | 0 | 0 | 0 | 0 | 1(20%) | 0 |
| **Exposure history** |  |  |  |  |  |  |
| Recently visited Wuhan | 0 | 1(50%) | 1(50%) | 0 | 0 | 0 |
| Contact with Wuhan residents | 2(100%) | 1(50%) | 0 | 4(66.7%) | 4(80%) | 3(100%) |
| Smoking history | 0 | 0 | 1(50%) | 2(33.3%) | 2(40%) | 0 |
| Mean latency (day) | 9 | 8.5 | 20.5 | 8.8 | 7 | 13.7 |
| **Comorbidities** |  |  |  |  |  |  |
| ARDS | 0 | 0 | 0 | 0 | 0 | 0 |
| Acute kidney injury | 0 | 0 | 0 | 0 | 0 | 0 |
| Myocardial injury | 0 | 0 | 0 | 1(16.7%) | 0 | 0 |
| Liver dysfunction | 0 | 0 | 0 | 0 | 0 | 0 |
| Urinary tract infection | 0 | 0 | 0 | 0 | 0 | 0 |
| Digestive system damage | 0 | 0 | 0 | 0 | 0 | 0 |
| **Abnormalities on chest CT** |  |  |  |  |  |  |
| Ground-glass opacity | 0 | 1(50%) | 1(50%) | 1(16.7%) | 1(20%) | 2(66.7%) |
| Bilateral patchy shadowing | 0 | 1(50%) | 0 | 0 | 1(20%) | 0 |
| No obvious abnormalities | 2(100%) | 0 | 0 | 4(66.7%) | 0 | 0 |
| **Laboratory findings** |  |  |  |  |  |  |
| White blood cell count | 4.66 | 3.9 | 7.105 | 5.9 | 6.42 | 5.19 |
| Median (IQR) | (3.81-5.5) |  | (5.11-9.1) | (4.11-6.27) | (6.68-9.4) | (5.17-8.05) |
| Neutrophil Percentage | -- | 46.8% | 62.15% | 63% | 62.5% | 69.2% |
| median(IQR) |  | (35.6-58.0%) | (60.1-64.2%) | (41.6-83.2%) | (60.2-67%) | (61.4-77.1%) |
| Lymphocyte count | 2.925 | 1.63 | 2.135 | 1.3 | 2.04 | 0.83 |
| Median (IQR) | (1.91-3.94) | (1.21-2.05) | (1.31-2.96) | (0.96-1.99) | (2.03-3) | (0.78-2.57) |
| **T cell detection** |  |  |  |  |  |  |
| CD4+ T cell count | 811 | 691 | 420 | 433 | 253 | 273 |
|  | (768-854) |  |  | (211-886) | (239-1251) | (188-1033) |
| CD8+ T cell count | 561 | 537 | -- | 370 | -- | -- |
|  |  |  |  | (159-516) |  |  |
| CD4+/CD8+ | 1.37 | 1.29 | -- | 1.33 | -- | -- |
|  |  |  |  | (1.13-2.40) |  |  |
| Platelet count, Median | 140.5 | 166 | 229 | 183 | 197 |  |
| (IQR) | (30-251) |  | (221-237) | (87-328) | (192-198) |  |
| Elevated C-reactive protein (>10 mg/L) | 0 | 1 | 0 | 1 | 1 | 0 |
|  |  | (50%) |  | (16.7%) | (20%) |  |
| Elevated Procalcitonin (>0.5ng/ml) | 1 | 0 | 0 | 1 | 0 | 0 |
|  | (50%) |  |  | (16.7%) |  |  |
| **Clinical outcome** |  |  |  |  |  |  |
| Recovery | 2(100%) | 2(100%) | 2(100%) | 6(100%) | 5(100%) | 3(100%) |
| Hospital length of stay | 16.5 | 10.5 | 16 | 12.2 | 11.6 | 19 |
| (days) | (14-19) | (8-13) | (7-25) | (10-15) | (6-15) | (14-22) |
| Death | 0 | 0 | 0 | 0 | 0 | 0 |

Data are median (IQR), n (%) or mean (SD). Some patients lack certain tests, or some patients' mean latency is unknown. White blood cell count ( $\times 10^9$ cells per L). Lymphocyte count ( $\times 10^9$ cells per L). Platelet count, $\times 10^9$ per L. ARDS=acute respiratory distress syndrome. Patients means people who have confirmed SARS-CoV-2 infection or are highly suspected of being infected.
\*--means the data were missing.

### 4. Clinical outcome of SARS-CoV-2 infected patients in Chongqing

With the extension of the epidemic, the mean latency of SARS-CoV-2 infection patients was prolonged (10.7 days vs 7 days), but the hospital length of stay was shortened (15.8 days VS 18 days). The mean latency for SARS-CoV-2 infected patient who came back from Wuhan was the longest (9.2 days), and the average hospital length of stay decreased by patients return from Wuhan, patients exposure to Wuhan population, and patients with no more contact (18.8 days, 15.9 days, 15.2 days). Suggesting that the mean latency of SARS-CoV-2 infection may increase with the increase of its transmission chain, and the hospital length of stay for patients with a large number of transmission chains is shortening, indicated that the toxicity of SARS-CoV-2 may be reducing in the process of transmission. Moreover, the mean latency of asymptomatic patients was the longest (10.5 days); in contrast, the hospital length of stay was the shortest (14.3 days). These results suggest that SARS-CoV-2 infection may increase the mean latency due to its cryptic nature. However, the hospital length of stay may be shorten due to decreased toxicity during transmission. In addition, the mean latency was long in children and older group(10.8 days, 11.3 days). Overall, the hospital length of stay increases with age. This suggests that in the process of epidemic prevention and control, special attention needs to be paid to children and elderly people. See Table 4 for the detailed data.

**Table 4.**
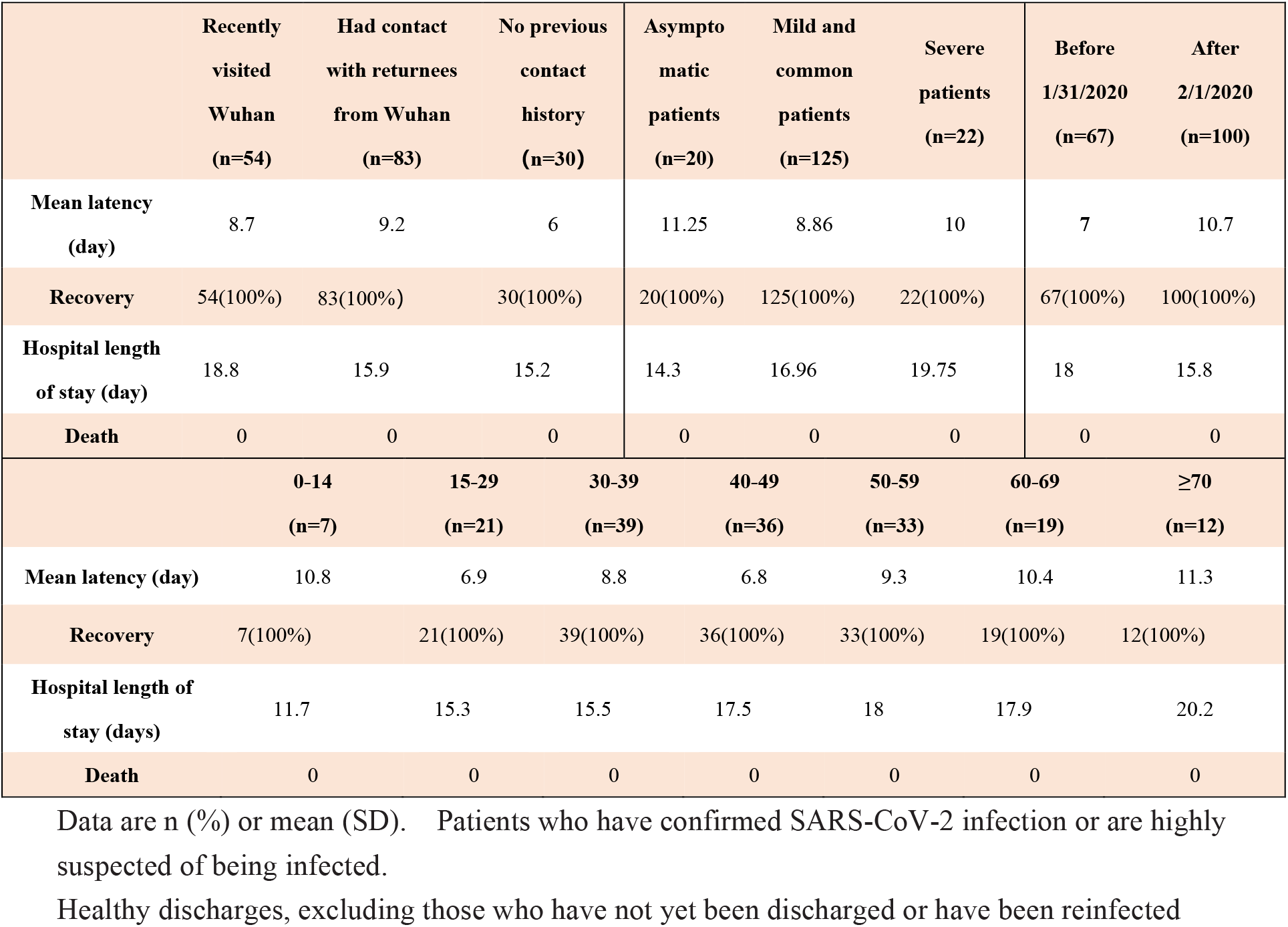
Developmental outcomes of SARS-CoV-2 infected patients.

|  | Recently<br>visited<br>Wuhan<br>(n=54) | Had contact<br>with returnees<br>from Wuhan<br>(n=83) | No previous<br>contact<br>history<br>(n=30) | Asympto<br>matic<br>patients<br>(n=20) | Mild and<br>common<br>patients<br>(n=125) | Severe<br>patients<br>(n=22) | Before<br>1/31/2020<br>(n=67) | After<br>2/1/2020<br>(n=100) |
| --- | --- | --- | --- | --- | --- | --- | --- | --- |
| **Mean latency**<br>**(day)** | 8.7 | 9.2 | 6 | 11.25 | 8.86 | 10 | 7 | 10.7 |
| **Recovery** | 54(100%) | 83(100%) | 30(100%) | 20(100%) | 125(100%) | 22(100%) | 67(100%) | 100(100%) |
| **Hospital length**<br>**of stay (day)** | 18.8 | 15.9 | 15.2 | 14.3 | 16.96 | 19.75 | 18 | 15.8 |
| **Death** | 0 | 0 | 0 | 0 | 0 | 0 | 0 | 0 |
|  | **0-14**<br>**(n=7)** | **15-29**<br>**(n=21)** | **30-39**<br>**(n=39)** | **40-49**<br>**(n=36)** | **50-59**<br>**(n=33)** | **60-69**<br>**(n=19)** | **≥70**<br>**(n=12)** |  |
| **Mean latency (day)** | 10.8 | 6.9 | 8.8 | 6.8 | 9.3 | 10.4 | 11.3 |  |
| **Recovery** | 7(100%) | 21(100%) | 39(100%) | 36(100%) | 33(100%) | 19(100%) | 12(100%) |  |
| **Hospital length of**<br>**stay (days)** | 11.7 | 15.3 | 15.5 | 17.5 | 18 | 17.9 | 20.2 |  |
| **Death** | 0 | 0 | 0 | 0 | 0 | 0 | 0 |  |

Data are n (%) or mean (SD). Patients who have confirmed SARS-CoV-2 infection or are highly suspected of being infected.
Healthy discharges, excluding those who have not yet been discharged or have been reinfected

## 5. Recurrence cases

### Case A (Recurrence of positive SARS-CoV-2 RNA in COVID-19)

A 34-year-old woman was admitted at 1.31(nonsevere), without comoebidities, had been to Wuhan 5 days ago. The CT scan revealed scattered infections in both lungs. White-cell count is 2.69× 10^9^ cells per L, Neutrophil ratio is 54.3%, Lymphocyte count is 0.96× 10^9^ cells per L, not admitted to the ICU. Nineteen days later, she was discharged healthy. Upon discharge, the CT scan of her discharge revealed improvement in the absorption of both lung lesions, White-cell count is 5.43× 10^9^ cells per L, Lymphocyte count is 1.21× 10^9^ cells per L.

### Case B (Recurrence of positive SARS-CoV-2 RNA in COVID-19)

A 44-year-old man was admitted at 2.04(nonsevere), without comoebidities, had not contacted Wuhan returnees or traveled to Wuhan. The CT scan revealed scattered infections in both lungs. White-cell count is 2.36× 10^9^ cells per L, Neutrophil ratio is 60.6%, Lymphocyte count is 0.6× 10^9^ cells per L, CD4^+^ T cell count is 196, CD8^+^ T cell count is 89, CD4^+^ /CD8^+^ is 2.22, C-reactive protein is 26.23 mg/L, Procalcitonin is normal, not admitted to the ICU. Eighteen days later, he was discharged healthy. Upon discharge, the CT scan revealed improvement in the absorption of both lung lesions, White-cell count is 5.43× 10^9^ cells per L, Lymphocyte count is 1.16× 10^9^ cells per L.

### Case C (Recurrence of positive SARS-CoV-2 RNA in COVID-19)

A 38-year-old man was admitted at 1.29(severe), without comoebidities, had contact with returnees from Wuhan 4 days ago. The CT scan revealed potential small patchy, flaky ground glass shadows on both lungs. White-cell count is 5.89× 10^9^ cells per L, Neutrophil ratio is 75.6%, Lymphocyte count is 2.58× 10^9^ cells per L, CD4^+^ T cell count is 252/ul, CD8^+^ T cell count is 305/ul, CD4^+^ /CD8^+^ is 0.83, C-reactive protein is 71.45 mg/L, Procalcitonin is 0.073 ng/ml, not admitted to the ICU. Twenty-nine days later, he was discharged healthy. Upon discharge, the CT scan revealed the lungs were scattered in stellate, patchy, grid-like ground glass stoves, and cord shadows. White-cell count is 5.77× 10^9^ cells per L, Lymphocyte count is 1.55× 10^9^ cells per L, CD4^+^ T cell count is 299/ul, CD8^+^ T cell count is 264/ul, CD4^+^ /CD8^+^ is 0.83.

## Discussion

Chongqing currently has a severe Covid-19 rate of 13.17% of the 167 patients and no one died. Of the 22 severe Covid-19 patients, 13 (59%) had a history of travel in Wuhan; and of the 167 SARS-CoV-2 infected patients, 54 (32.33%) had a history of travel in Wuhan. Therefore, Chongqing ‘s epidemic situation presents typical characteristics of imported cases. The clinical characteristics, radiologic and laboratory indexes and clinical outcomes of these patients with severe Covid-19 are consistent with previous research reports, and only 3 patients (13.64%) had comorbidities of liver dysfunction, which was lower than the previously reported complication rate of 25.43%^9^.

WHO has positioned SARS-CoV-2 as a global pandemic, and asymptomatic patients with SARS-CoV-2 infection gradually appear^3^. A recent research report pointed out that a small number of asymptomatic patients with SARS-CoV-2 infection were observed in the imported population from Wuhan^10^. Our results show that 2 out of 54 imported cases are asymptomatic patients in Chongqing, accounted for 3.7%. The proportion of asymptomatic patients among people infected with SARS-CoV-2 due to contact with imported populations was higher, reaching 18.1%. Although SARS-CoV-2 infection has been reported in children aged 0-14 years^11^, we found 2 children were asymptomatic patients with SARS-CoV-2 infection for the first time, accounting for 28.6% of infected children. 3 cases of asymptomatic SARS-CoV-2 infection in elderly group, accounting for 25% of the infections in this age group. The vast majority (66.67%) of asymptomatic patients over 70 years of age are accompanied by imaging changes, indicating that this part of the population can be screened by imaging studies. In children and young adults with asymptomatic SARS-CoV-2 infection, although there are changes such as bronchial thickening, there are not many typical clinical changes. As the epidemic develops, the proportion of asymptomatic patients is getting higher and higher (up to 19% in the onset after February 1^st^, 2020). The mean latency of asymptomatic patients with SARS-CoV-2 infection was prolonged (10.5 days), but the hospital length of stay was shortened (14.3 days).

All countries have adopted isolation measures for imported populations from areas or countries with severe SARS-CoV-2 infection, and treated patients with symptoms who have a history of contact with this imported population^1^. However, a large number of asymptomatic patients with contact history with imported cases failed to be included in the focus surveillance. Our study shows for the first time that asymptomatic patients with SARS-CoV-2 infection have a contact history with imported case account for a large proportion. Therefore, the next stage of SARS-CoV-2 prevention and control needs to focus on the screening of asymptomatic patients in the community with a history of contact with the imported population, especially for children and the elderly. Promoting the worldwide prevention and control of SARS-CoV-2.

Limitations of this study: (1) Because some SARS-CoV-2 infected patients have been discharged from hospitals, and the early epidemic situation is severe, medical workers are devoted to fighting the epidemic, resulting in the loss of some patients’ files. (2) 167 Case data we used were from the Chongqing Public Health Medical Center, and the number of asymptomatic infections is 20, which is relatively small. More case data of asymptomatic patients are needed to confirm our results. (3) Due to the huge workload of medical workers, patients with pneumonia infected by virus complex bacteria are not distinguished. (4) Our data is artificially calculated and not automatically generated by the system, so it has a certain degree of subjectivity.

## Data Availability

After publication, the data will be made available to others on reasonable requests to the corresponding author. A proposal with detailed description of study objectives and statistical analysis plan will be needed for evaluation of the reasonability of requests. Additional materials might also be required during the process of evaluation. Deidentified participant data will be provided after approval from the corresponding author and Chongqing Public Health Medical Center.

## Contributors

Yang Tao, Panke Cheng and Wen Chen collected the epidemiological and clinical data and drafted the manuscript. Peng Wan, Yaokai Chen, Guodan Yuan, Junjie Chen collected and summarised all data. Da Huo, Ge Guan, Dayu Sun, Ju Tan and Guanyuan Yang draw figures and tables. Weng Zeng and Chuhong Zhu designed the clinical study, analyzed all data and revised the final manuscript.

## Acknowledgments

We thank all patients and their families involved in the study. We thank all doctors and nurses in Chongqing Public Health Medical Center. This work was supported by the National Key Research and Development Program (No.2016YFC1101100) and National Science Fund for Distinguished Young Scholars (No. 31625011).

---

**Table S1.** Variable of mild and common patients at different ages

|  | 0-14(n=5) | 15-29(n=19) | 30-39(n=28) | 40-49(n=30) | 50-59(n=26) | 60-69(n=13) | ≥70(n=4) |
| --- | --- | --- | --- | --- | --- | --- | --- |
| **All patients before 1/31/2020** | 2(40%) | 11(57.9%) | 18(64.3%) | 6(20%) | 9(34.6%) | 5(38.5%) | 1(25%) |
| **Exposure history** |  |  |  |  |  |  |  |
| **Recently visited Wuhan** | 1(20%) | 6(31.6%) | 16(57.1%) | 5(16.7%) | 8(30.8%) | 4(30.8%) | 1(25%) |
| **Contact with Wuhan residents** | 4(80%) | 7(36.8%) | 8(28.6%) | 18(60%) | 7(26.9%) | 9(69.2%) | 3(75%) |
| **Smoking history** | 0 | 1(5.3%) | 5(17.9%) | 4(13.3%) | 2(7.7%) | 2(15.4%) | 0 |
| **Mean latency (day)** | 11.7 | 6.57 | 7.2 | 6.3 | 9.6 | 10.4 | 10.25 |
| **Clinical symptoms** |  |  |  |  |  |  |  |
| Fever | 1(20%) | 13(68.4%) | 21(75%) | 14(46.7%) | 15(57.7%) | 6(46.2%) | 2(50%) |
| Cough | 5(100%) | 12(63.2%) | 20(71.4%) | 22(73.3%) | 14(53.8%) | 10(76.9%) | 3(75%) |
| Shortness of breath | 0 | 1(5.3%) | 7(25%) | 2(6.7%) | 5(19.2%) | 3(23.1%) | 1(25%) |
| Fatigue | 0 | 2(19.5%) | 7(25%) | 7(23.3%) | 5(19.2%) | 5(38.5%) | 0 |
| **Comorbidities** |  |  |  |  |  |  |  |
| ARDS |  |  | 0 | 0 | 0 | 0 | 0 |
| Acute kidney injury | 0 | 0 | 0 | 0 | 0 | 0 | 0 |
| Myocardial injury | 0 | 0 | 0 | 0 | 0 | 0 | 0 |
| Liver dysfunction | 0 | 1(5.3%) | 1(3.6%) | 2(6.7%) | 1(3.8%) | 1(7.7%) | 0 |
| Urinary infection | 0 | 0 | 0 | 0 | 0 | 0 | 0 |
| Digestive system damage | 0 | 0 | 0 | 0 | 0 | 0 | 0 |
| **Abnormalities on chest CT** |  |  |  |  |  |  |  |
| Ground-glass opacity | 0 | 6(31.6%) | 16(57.1%) | 8(26.7%) | 8(30.8%) | 6(46.2%) | 1(25%) |
| Bilateral patchy shadowing | 0 | 3(15.8%) | 4(14.3%) | 4(13.3%) | 5(19.2%) | 5(38.5%) | 1(25%) |
| No obvious abnormalities | 1(20%) | 2(10.5%) | 0 | 0 | 0 | 0 | 0 |
| **Laboratory findings** |  |  |  |  |  |  |  |
| White-cell count Median | 6.62 | 4.36 | 4.31 | 5.01 | 5.25 | 5.46 | 4.03 |
| (IQR) | (4.14-9.1) | (2.7-9.67) | (2.62-18.72) | (2.36-10.53) | (2.99-8.79) | (2.24-6.93) | (3.83-9.06) |
| Neutrophil Percentage | 48.85% | 43.97% | 52.25% | 61.79% | 75.6% | 69.80% | 72.3% |
| median(IQR) | (41.50-56.20%) | (19.15-61.9) | (36.86%-89.2%) | (47.62%-92.12%) | (63.3-78.5%) | (36.7-79.9%) | (58.7-86%) |
| Lymphocyte count Median | 2.2 | 1.605 | 1.32 | 1.05 | 1.05 | 1.13 | 0.8 |
| (IQR) | (1.2-3.2) | (0.95-3.17) | (0.69-2.26) | (0.6-2.32) | (0.41-2.31) | (0.64-2.24) | (0.78-1.19) |
| T cell detection | 3209 | 1086 | 867.4 | 815 | 765 | 873.4 | 548 |
|  | (3209) | (656-2250) | (406-1527) | (369-1620) | (350-1374) | (373-1066) | (473-588) |
| CD4+ T cell count | 869 | 459 | 419.4 | 430.5 | 423.7 | 495.3 | 308.3 |
|  | (347-1770) | (229-1209) | (174-886) | (185-804) | (191-774) | (205-848) | (249-371) |
| CD8+ T cell count | 790 | 526 | 337 | 287.6 | 278.3 | 303 | 169.5 |
|  | (667-1439) | (231-991) | (158-628) | (89-765) | (54-568) | (121-517) | (152-187) |
| CD4+/CD8+ | 1.1(0.52-1.23) | 0.965 | 1.36 | 1.60 | 2.02 | 1.67 | 1.99 |

|  |  | (0.54-2.46) | (0.79-1.94) | (0.86-2.84) | (0.54-3.73) | (1.25-1.99) | (1.98-2) |
| --- | --- | --- | --- | --- | --- | --- | --- |
| Platelet count, Median | 241 | 147 | 186 | 140.5 | 173 | 233 | 200 |
| (IQR) | (241) | (105-341) | (79-455) | (94-350) | (67-501) | (68-331) | (158-424) |
| Elevated C-reactive protein<br>($>10$ mg/L) | 1(20%) | 1(5.3%) | 9(32.1%) | 11(36.7%) | 5(19.2%) | 4(30.8%) | 2(50%) |
| Elevated Procalcitonin<br>($>0.5$ng/ml) | 0 | 0 | 2(7.1%) | 3(10%) | 2(7.7%) | 4(30.8%) | 0 |
| **Clinical outcome** |  |  |  |  |  |  |  |
| Recovery | 5(100%) | 19(100%) | 28(100%) | 30(100%) | 26(100%) | 13(100%) | 4(100%) |
| Hospital length of stay<br>(days) | 9.8 | 15.83 | 15.07 | 19.55 | 17.25 | 19.25 | 22 |
| Death | 0 | 0 | 0 | 0 | 0 | 0 | 0 |

Data are median (IQR), n (%) or mean (SD). Some patients lack certain tests, or some patients' mean latency is unknown. White blood cell count ( $\times 10^9$ cells per L). Lymphocyte count ( $\times 10^9$ cells per L) . Platelet count, $\times 10^9$ per L. ARDS=acute respiratory distress syndrome. Patients means people who have confirmed SARS-CoV-2 infection or are highly suspected of being infected.

